# Efficacy and Safety of Lopinavir/Ritonavir for Treatment of COVID-19: A Systematic Review and Meta-Analysis

**DOI:** 10.1101/2020.06.16.20133298

**Authors:** Mansour Tobaiqy, Saad Alhumaid, Abbas Al Mutair

## Abstract

**Background:** Since the emergence of COVID-19, the world has been desperate to find effective therapeutics and vaccinations to treat hundreds of thousands of affected patients and to suppress the spread of this global pandemic. Lopinavir-ritonavir (LPV/RTV) is an HIV antiviral combination that has been considered for treatment of this disease.

**Aim of the study:** This systematic review and meta-analysis aimed to assess the efficacy and safety of lopinavir/ritonavir in COVID-19 patients in the extant published research. A systematic review protocol was developed based on PRISMA-P and the PRISMA statement. Published English and non-English articles written since December 1, 2019 were selected for review from 8 electronic databases.

Readily accessible full articles (cohort studies and clinical trials) which specifically mentioned lopinavir/ritonavir and patients with lab-confirmed SARS-CoV-2 or COVOID-19 of any age were included. Three researchers separately evaluated the bias in the reported articles. We conducted a systematic review and meta‐analysis with the objective of evaluation of the safety and efficacy of LPV/RTV alone or in combination with other drugs with regard to time to becoming PCR negative, time to body temperature normalization and cough relief, radiological progression, and safety. Review Manager (RevMan) was used to conduct all statistical analyses and generate the forest plots. Meta-analyses were performed using the Mantel Hazel method or the inverse variance method for dichotomous data and continuous data respectively.

**Results:** Non-duplicate articles (n=76) were evaluated for possible inclusion. A consensus was reached to select 29 articles for full-text screening, only 11 articles comprised 1,192 patients were included in this study, and six of which were included for meta-analysis.

In terms of virological cure (PCR negative), three studies reported less time in days to achieve a virological cure for LPV/RTV arm relative to no antiviral therapy (conventional) (mean difference = −0.81 day; 95% CI, −4.44 to 2.81; *P* = 0.007, *I*^*2*^ = 80%). However, the overall effect was not significant (*P* = 0.66). When comparing LPV/RTV arm to umifenovir arm, a favorable affect was observed for umifenovir arm, but not statically significant (mean difference = 0.95 day; 95% CI, −1.11 to 3.01; *P* = 0.09, *I*^*2*^ = 58%).

In terms of time to body normalization and cough relief (clinical cure), two studies reported on time to temperature normalization with no significant effect of LPV/RTV (n = 93) versus umifenovir (n = 71) arm), (OR = 0.87 day; 95% CI, 0.42 to 1.78; (*P* = 0.70), I2 = 0%), or alleviation of cough duration (p = 0.69).

In terms of CT evidence of radiological progression of pneumonia/lung damage, treatment with lopinavir/ritonavir resulted in no significant decrease in the radiological progression (OR = 0.80; 95% CI, 0.42 to 1.54; *P* = 0.59, *I*^*2*^ = 81%), In terms of safety, a greater number of adverse events were reported for lopinavir/ritonavir (n=45) relative to the umifenovir arm (n=14) and conventional treatments (n=10), *P* = 0.004, 0,0007, respectively

**Conclusions:** The small number of studies included in this systematic review and meta-analysis study did not reveal any statistically significant advantage in efficacy of lopinavir-ritonavir in COVID-19 patients, over conventional or other antiviral treatments. This result might not reflect the actual evidence.

## Introduction

Since the emergence of an unknown viral infection in China in December 2019 and following the identification of this infection as 2019-new coronavirus disease (2019-nCoV, also known as COVID-19), caused by severe acute respiratory syndrome coronavirus 2 (SARS-CoV-2) [1], the world is desperate to find effective therapeutics and vaccinations to treat hundred thousand of affected patients and to reduce the spread of this global pandemic [2].

As June 2, there are 1104 registered <u>clinical trials of COVID-19</u> therapeutics or vaccinations worldwide that either ongoing or recruiting patients; however, no drug or vaccine has officially been approved for COVID-19 [2, 3]. These trials have produced mixed and conflicting results of positive or negative outcomes and inclusive evidence of efficacy or safety, that render the suspension of some trials inevitable, as in the hydroxychloroquine trials, which was suggested by World Health Organization (WHO) in light of safety concerns [4]. This decision was reversed on June 3, 2020 [5] following a retraction of the research article by Lancet as certain authors were not granted access to the underlying data [6].

Lopinavir-ritonavir is a protease inhibitor and nucleoside analogue combination used for human immunodeficiency virus (HIV-1), and is also considered a potential treatment for COVID-19 [7], as its therapeutic value in the treatment of COVID-19 emerged from in-vitro studies that demonstrated inhibition of several viral corona respiratory illnesses, including severe acute respiratory syndrome (SARS-CoV), and Middle East Respiratory Syndrome (MERS) [8. 9, 10].

Lopinavir (LPV) is an aspartic acid protease inhibitor of HIV, where inhibition of proteases enzymes is essential for the intervening of the viral infectious cycle, and is co‐formulated with ritonavir to boost the pharmacokinetic activity and half‐life of lopinavir through inhibition of Cytochromes P450, providing adequate suppression of viral load and constant improvements in CD4+ cell counts, as demonstrated in randomized trials in naïve and experienced adults and children HIV patients [7].

Lopinavir/ritonavir is available as a single‐tablet formulation (Kaletra^®^) in dosage strengths of 400/100 mg or 200/100 mg, and in clinical trials, this combination reduced rates of acute respiratory distress syndrome (ARDS) or death compared to supportive care or ribavirin alone in a matched cohort group during the early phase of viral acquisition [8].

Lopinavir-ritonavir is being examined in several international clinical trials, including the RECOVERY trial and SOLIDARITY WHO trial [11], but did not gain authorization to be used emergently in the current pandemic in the USA by the Food and Drug Administration (FDA), which has approved only three pharmacologically different therapeutics for treatments of COVID-19, including antibiotic-hydroxychloroquine, immunotherapy-convalescent plasma therapy, and antiviral-remdesivir [2, 11].

Among the clinical trials that did not find positive results for lopinavir-ritonavir, a study conducted by Bin Cao et al. and published in New England Journal of Medicine [12] revealed that treatment with lopinavir–ritonavir was not associated with clinical improvement beyond standard care or reduction in mortality rate at 28 days in hospitalized adult patients with severe COVID-19 [12].

To date, lopinavir/ritonavir combination is available in most countries’ therapeutics guidelines including USA [13], Saudi Arabia [14], and Ireland [15], which means that the medicine has tenable evidence of efficacy; however, considering early negative and conflicting results have emerged [12], there is a need to assess the efficacy and safety of this COVID-19 treatment in a systematic manner.

### Aim of the study

This systematic review and meta-analysis aim to assess the efficacy and safety of lopinavir/ritonavir in COVID-19 patients in published research.

## Methods

This systematic review was conducted with reference to the basics of Cochrane Handbook for Systematic Reviews of Interventions (16), described as stated by the Preferred Reporting Items for Systematic reviews and Meta-Analysis (PRISMA) statement [17, 18].

### Search strategy and selection criteria

A systematic review protocol was developed based on PRISMA-P and the PRISMA statement. Published articles from December 1, 2019 to May 22, 2020 were selected for review from 8 electronic databases (PubMed, CINAHL, Embase, medRxiv, Proquest, Wiley online library, Medline, and Nature).

The focus of the review was lopinavir/ritonavir treatment in COVID-19 patients. The primary outcome was the efficacy of lopinavir/ritonavir in COVID-19 patients. The secondary outcome was adverse events associated with its use.

### Inclusion criteria

Readily accessible peer-reviewed full articles, observational cohort studies, and clinical trials were included.

### Participants

Patients with lab-confirmed COVOID-19 of any age were included.

### Intervention

The interventions were lopinavir/ritonavir versus a control of either no antiviral therapy (conventional therapy), standard therapy, or lopinavir/ritonavir with other antiviral agents.

### Objectives

A. Virological cure on day 7 after initiation of therapy (+ve to −ve PCR: non-detection of SARS-CoV-2 in nasopharyngeal swab).
B. Clinical cure (time to body temperature normalization and time to cough relief).
C. Radiological progression during drug treatment.
D. Safety and tolerability of lopinavir/ritonavir.

### Comparisons

A. lopinavir/ritonavir vs no antiviral therapy (conventional therapy)/control.
B. lopinavir/ritonavir in combination of other agents versus conventional therapy/control.

### Searching keywords

The search keywords included 2019-nCoV, 2019 novel coronavirus, COVID-19, coronavirus disease 2019, SARS-COV-2, lopinavir, ritonavir, combination, kaletra, treatment, efficacy, clinical trial, cohort, retrospective, and prospective.

### Exclusion criteria

Types of articles that were excluded included duplicate articles, editorials, reviews, case reports, and letters to editors.

Any research articles that did not include data on lopinavir/ritonavir use, did not include control patients’ group, or reported combined use of lopinavir/ritonavir with other antiviral medications were also excluded.

### Data extraction and analysis

Two reviewers (MT and SA) independently screened the titles with abstracts using the selection criteria. For relevant articles, full texts were obtained for further evaluation. Disagreements between the two reviewers after full text screening were reconciled via consensus by a third reviewer (AA) [19].

Inclusions and exclusions were recorded following PRISMA guidelines presented in the form of a PRISMA flow diagram and detailed reasons recorded for exclusion. Articles were categorized as clinical trials or cohort studies. The following data were extracted from selected studies: authors; publication year; study location; study design and setting; sample size, age, and gender; details of study intervention and control therapies in addition to data on adverse events and treatment outcomes; assessment of study risk of bias; and remarks on noticeable findings.

### Risk of biased evaluation of included studies

The quality assessment of the studies was undertaken based on the revised Cochrane risk of bias tool (RoB 2.0) for randomized controlled studies [20]. ROBINS-I tool was used to asses non-randomized interventional studies [21], and Newcastle Ottawa Scale for observational cohort studies [22]. Critical appraisal checklists appropriate to each study design were applied and checked by the third-team member.

Three investigators (MT, SA, and AA) separately evaluated the possibility of bias using these tools. Publication bias was not evaluated by funnel plot as there was only three studies which were included in the meta-analysis part of the study.

### Assessment of heterogeneity

Statistical heterogeneity was evaluated using the *χ*^*2*^ test and *I*^*2*^ statistics (17, 18). An *I*^*2*^ value of 0 to <40% was not considered as significant, 30% to 60% was regarded as moderate heterogeneity, 50% to 90% was considered substantial heterogeneity, and 75% to 100% was considered significant heterogeneity.

### Statistical analysis

Because all of the data were continuous and dichotomous data, we used either odds ratio (OR) or mean difference for estimating the point estimate, along with a 95% confidence interval (CI). In the absence of significant clinical heterogeneity, we performed the meta-analysis using the Mantel Hazel method or inverse variance method for dichotomous data and continuous data respectively. Employing a conservative approach, a random effects model was used, which produces wider CIs than a fixed effect model [16]. Review Manager (Version 5.3, Oxford, UK; The Cochrane Collaboration, 2014) was used to conduct all statistical analyses and generate the forest plots [18].

## Results

A total of 8 literature databases were screened and 76 non-duplicate articles was identified, which were evaluated for possible inclusion using titles and abstracts. Out of these, 29 articles were selected for full-text screening and finally, eleven articles (total participants= 1,192) were included in the systematic review and six articles were included in the meta-analysis. 18 articles were excluded following full-text screening (reasons: review= 5, study with no relative data= 6, LPV/RTV use data not available= 2, no control patients in the study= 1, combined LPV/RTV use with other antiviral therapies/other medications data= 2, no extractable data= 2). The PRISMA chart for the included studies is displayed in Figure 1. The details of the included studies are depicted in Table 1. Among these, two articles were in preprint versions [23, 24].

**Table 1:**
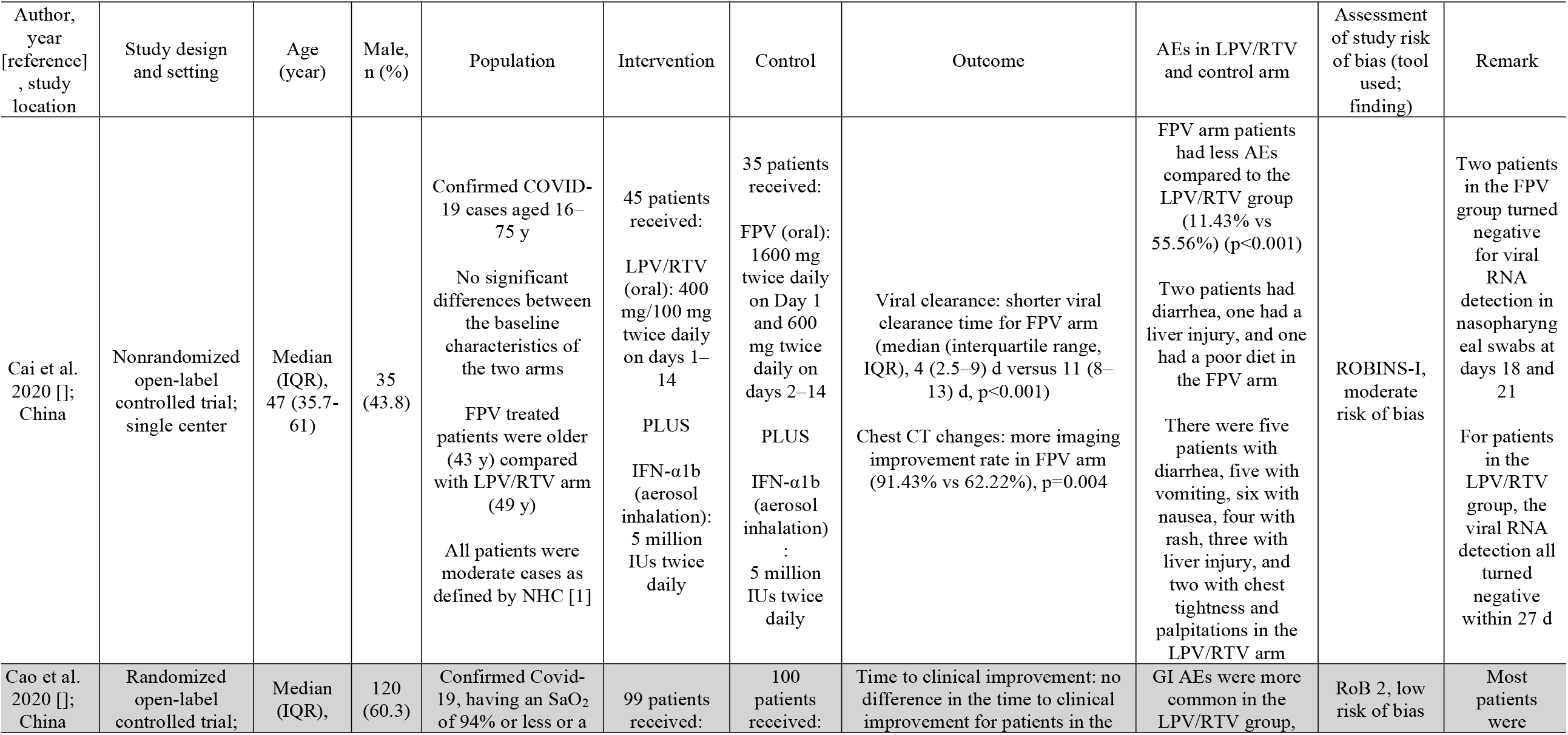
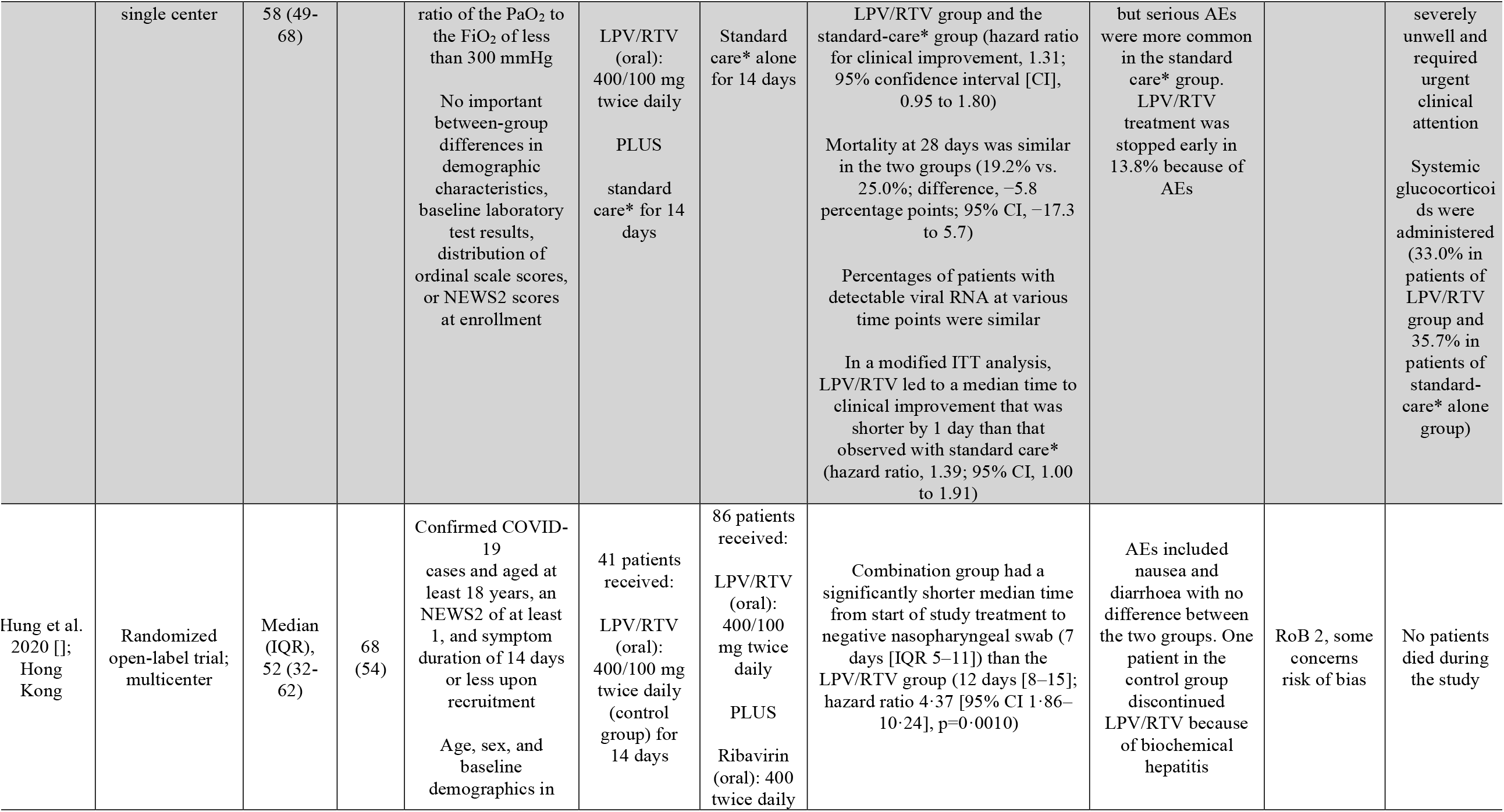
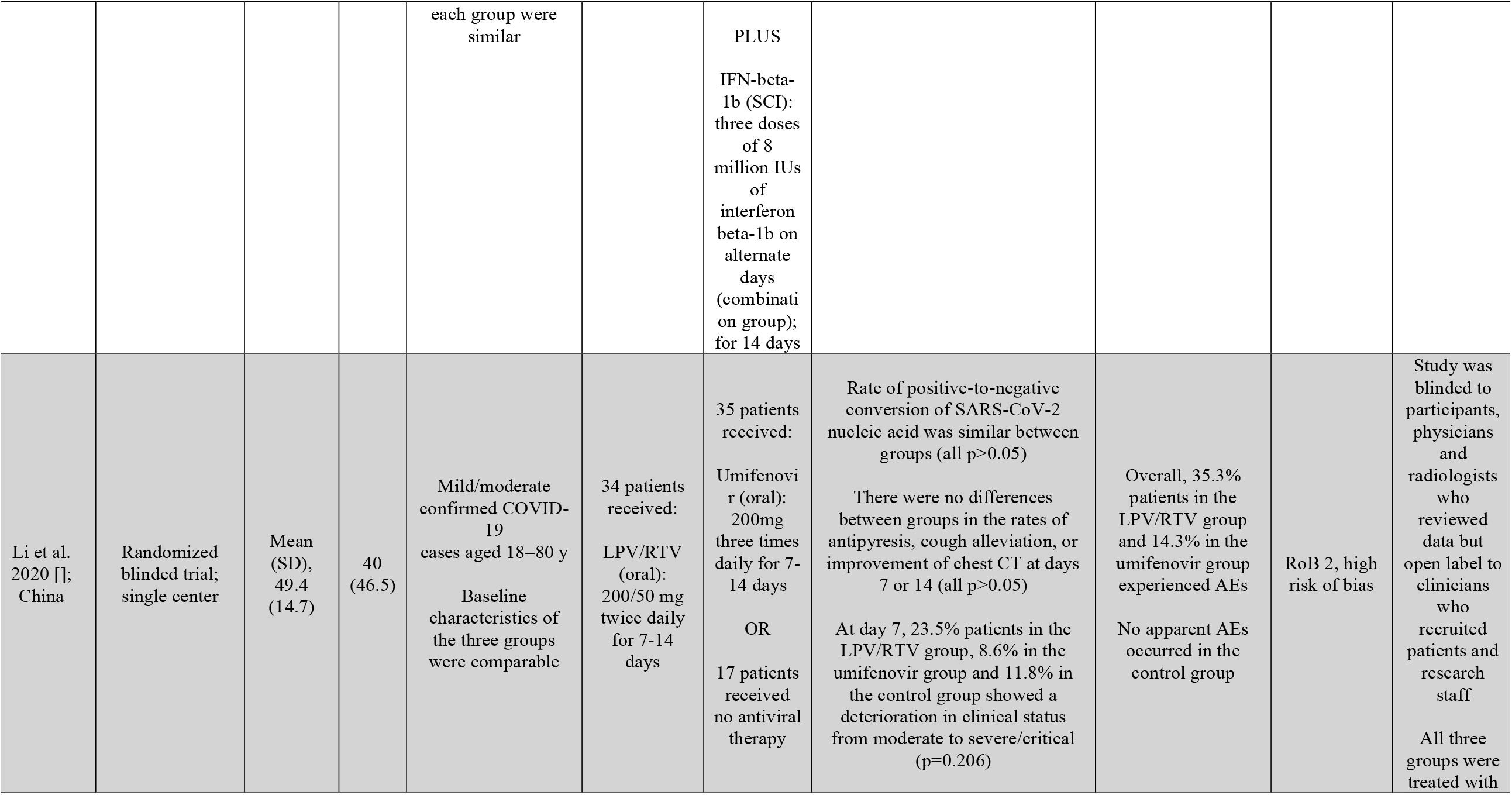
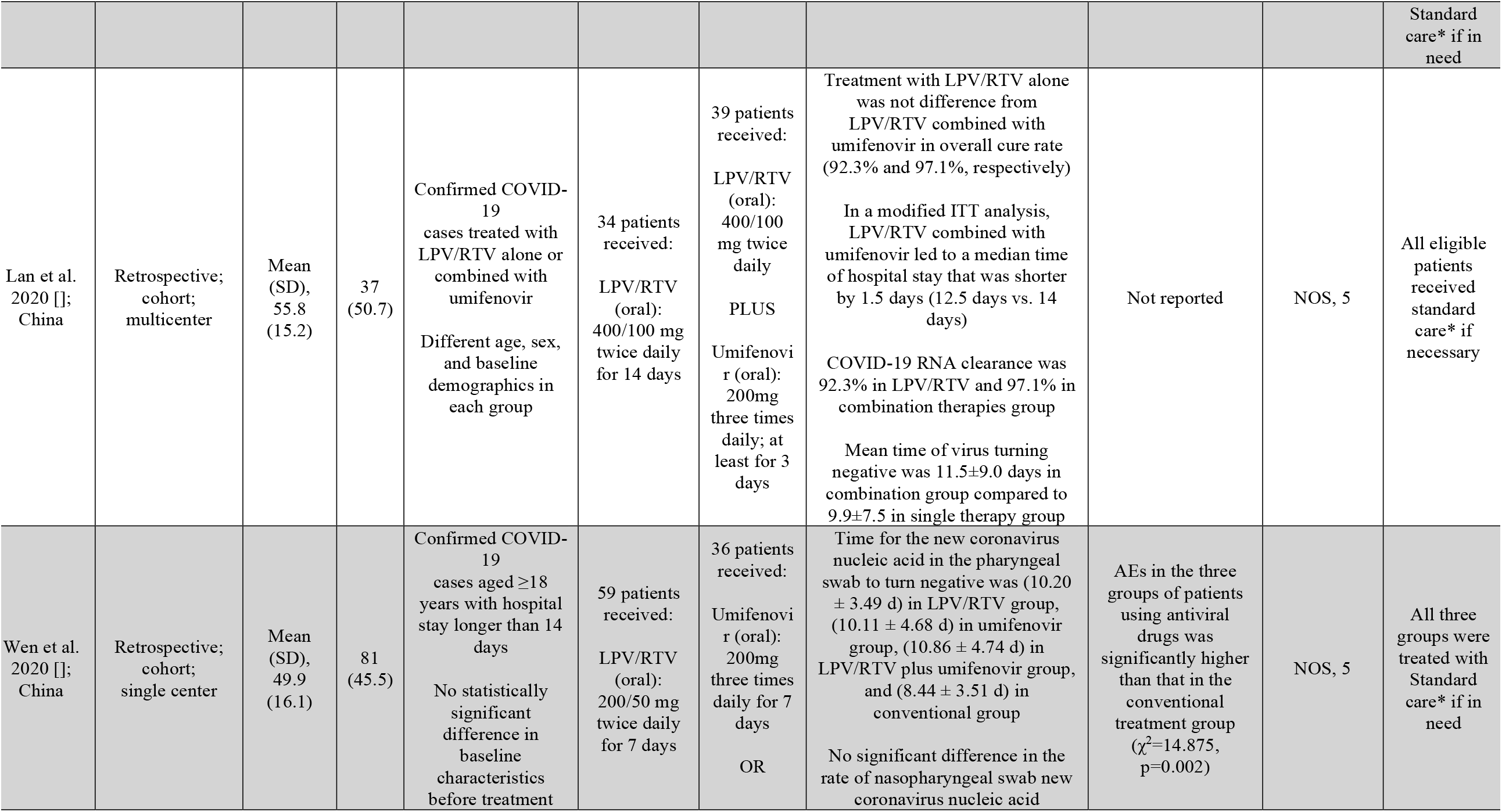
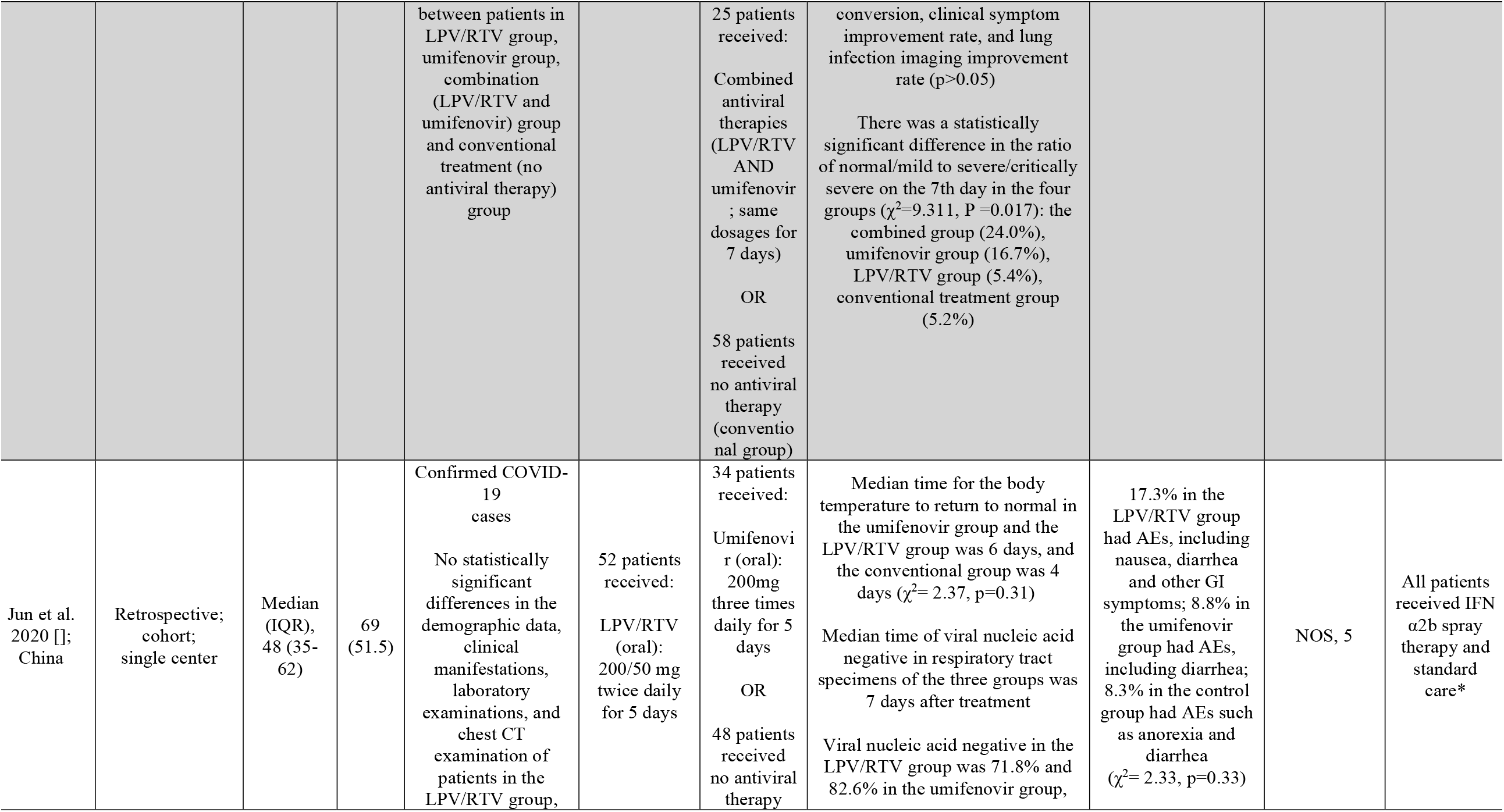
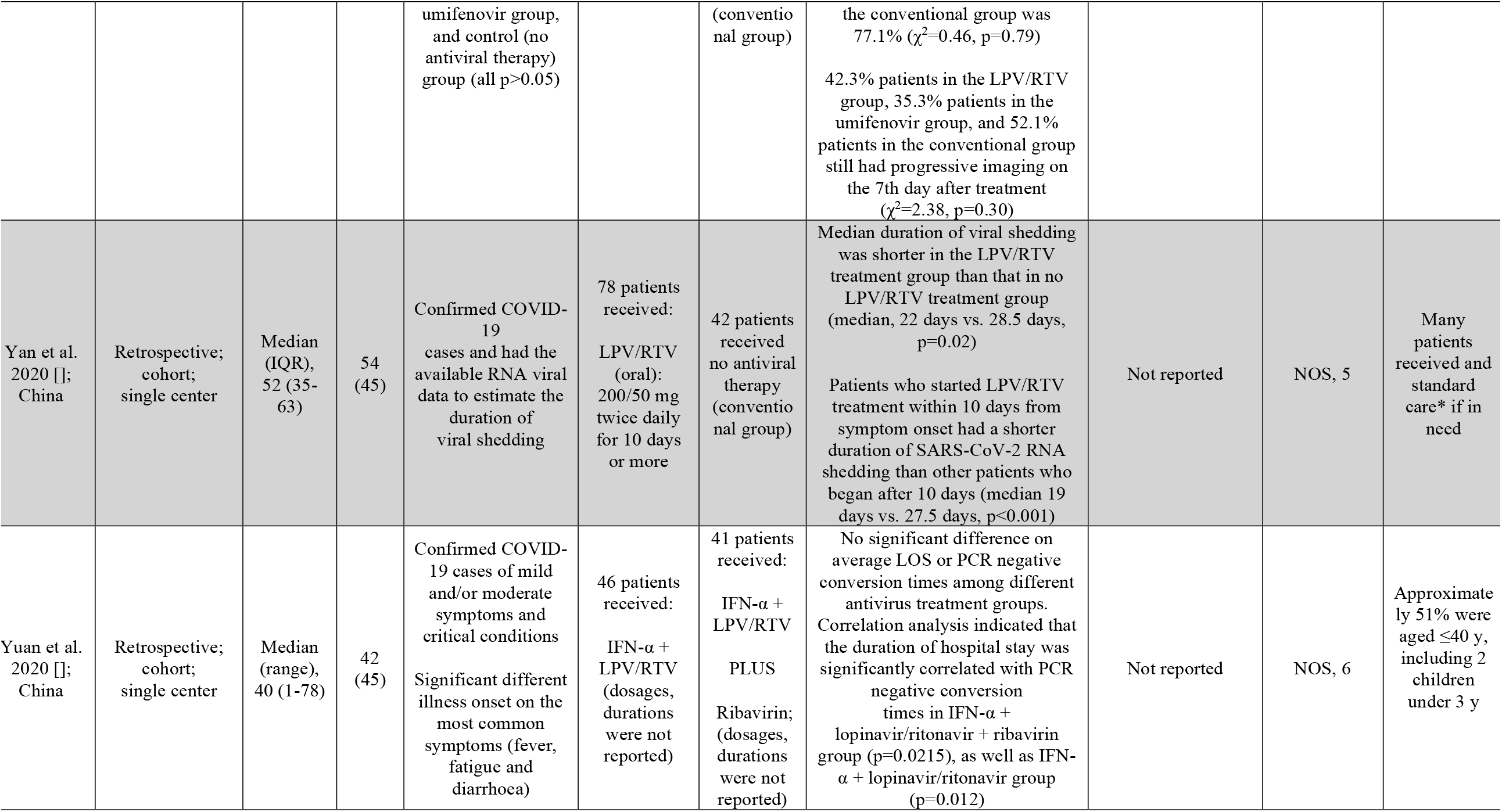
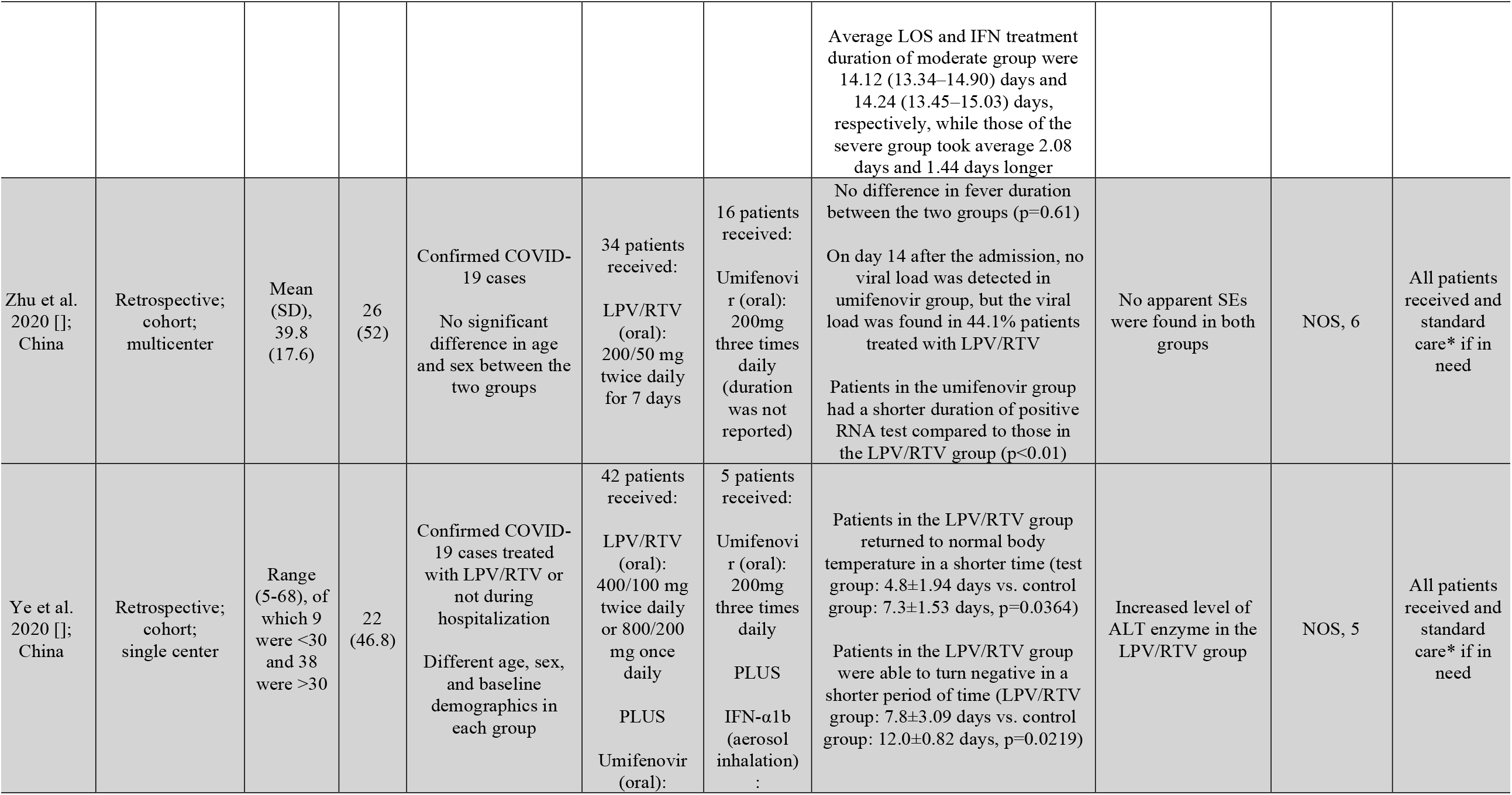
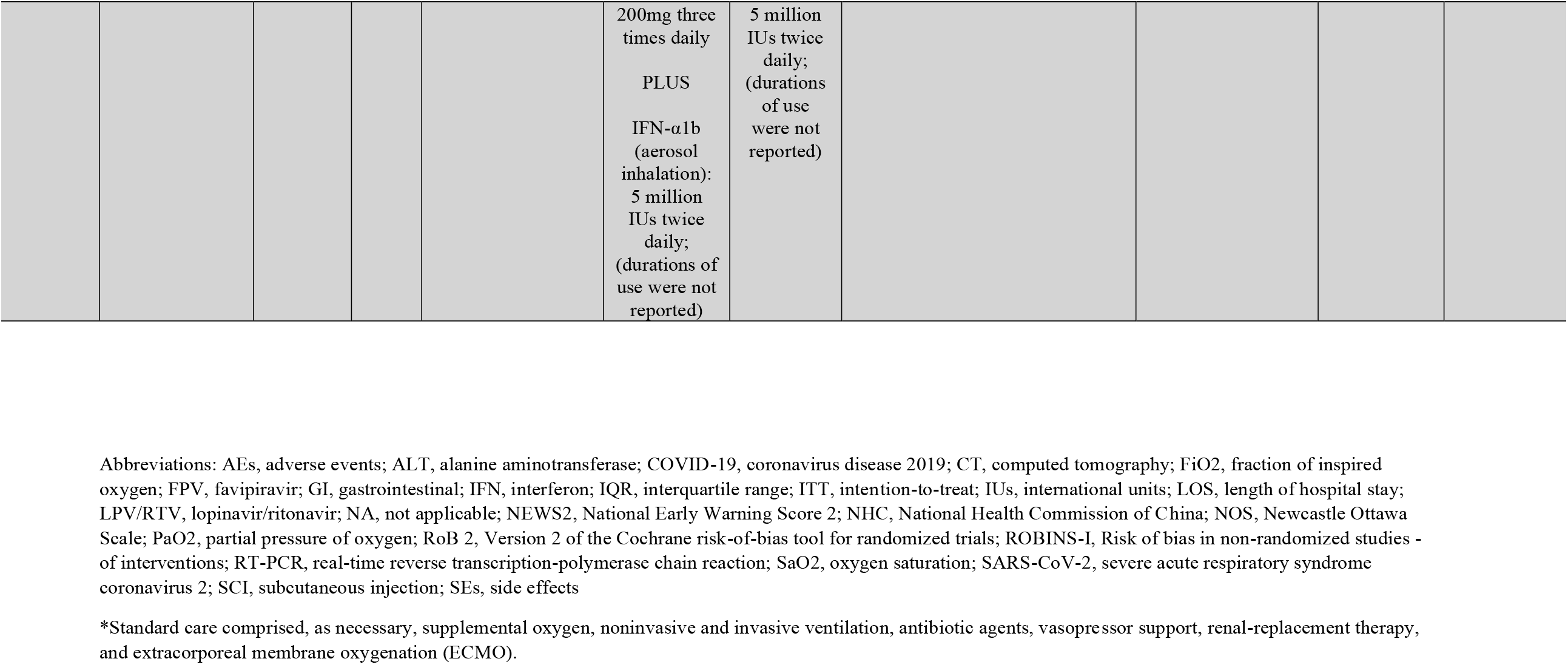
Data extracted from included papers (n=11)

| Author, year [reference], study location | Study design and setting | Age (year) | Male, n (%) | Population | Intervention | Control | Outcome | AEs in LPV/RTV and control arm | Assessment of study risk of bias (tool used; finding) | Remark |
| --- | --- | --- | --- | --- | --- | --- | --- | --- | --- | --- |
| Cai et al. 2020 [1]; China | Nonrandomized open-label controlled trial; single center | Median (IQR), 47 (35.7-61) | 35 (43.8) | Confirmed COVID-19 cases aged 16–75 y<br>No significant differences between the baseline characteristics of the two arms<br>FPV treated patients were older (43 y) compared with LPV/RTV arm (49 y)<br>All patients were moderate cases as defined by NHC [1] | 45 patients received:<br>LPV/RTV (oral): 400 mg/100 mg twice daily on days 1–14<br>PLUS<br>IFN-α1b (aerosol inhalation): 5 million IUs twice daily | 35 patients received:<br>FPV (oral): 1600 mg twice daily on Day 1 and 600 mg twice daily on days 2–14<br>PLUS<br>IFN-α1b (aerosol inhalation): 5 million IUs twice daily | Viral clearance: shorter viral clearance time for FPV arm (median (interquartile range, IQR), 4 (2.5–9) d versus 11 (8–13) d, p<0.001)<br>Chest CT changes: more imaging improvement rate in FPV arm (91.43% vs 62.22%), p=0.004 | FPV arm patients had less AEs compared to the LPV/RTV group (11.43% vs 55.56%) (p<0.001)<br>Two patients had diarrhea, one had a liver injury, and one had a poor diet in the FPV arm<br>There were five patients with diarrhea, five with vomiting, six with nausea, four with rash, three with liver injury, and two with chest tightness and palpitations in the LPV/RTV arm | ROBINS-I, moderate risk of bias | Two patients in the FPV group turned negative for viral RNA detection in nasopharyngeal swabs at days 18 and 21<br>For patients in the LPV/RTV group, the viral RNA detection all turned negative within 27 d |
| Cao et al. 2020 [2]; China | Randomized open-label controlled trial; | Median (IQR), | 120 (60.3) | Confirmed Covid-19, having an SaO<sub>2</sub> of 94% or less or a | 99 patients received: | 100 patients received: | Time to clinical improvement: no difference in the time to clinical improvement for patients in the | GI AEs were more common in the LPV/RTV group, | RoB 2, low risk of bias | Most patients were |
|  | single center | 58 (49-68) |  | ratio of the PaO<sub>2</sub> to the FiO<sub>2</sub> of less than 300 mmHg<br>No important between-group differences in demographic characteristics, baseline laboratory test results, distribution of ordinal scale scores, or NEWS2 scores at enrollment | LPV/RTV (oral): 400/100 mg twice daily<br>PLUS<br>standard care* for 14 days | Standard care* alone for 14 days | LPV/RTV group and the standard-care* group (hazard ratio for clinical improvement, 1.31; 95% confidence interval [CI], 0.95 to 1.80)<br>Mortality at 28 days was similar in the two groups (19.2% vs. 25.0%; difference, -5.8 percentage points; 95% CI, -17.3 to 5.7)<br>Percentages of patients with detectable viral RNA at various time points were similar<br>In a modified ITT analysis, LPV/RTV led to a median time to clinical improvement that was shorter by 1 day than that observed with standard care* (hazard ratio, 1.39; 95% CI, 1.00 to 1.91) | but serious AEs were more common in the standard care* group. LPV/RTV treatment was stopped early in 13.8% because of AEs |  | severely unwell and required urgent clinical attention<br>Systemic glucocorticoids were administered (33.0% in patients of LPV/RTV group and 35.7% in patients of standard-care* alone group) |
| Hung et al. 2020 []; Hong Kong | Randomized open-label trial; multicenter | Median (IQR), 52 (32-62) | 68 (54) | Confirmed COVID-19 cases and aged at least 18 years, an NEWS2 of at least 1, and symptom duration of 14 days or less upon recruitment<br>Age, sex, and baseline demographics in | 41 patients received:<br>LPV/RTV (oral): 400/100 mg twice daily (control group) for 14 days | 86 patients received:<br>LPV/RTV (oral): 400/100 mg twice daily<br>PLUS<br>Ribavirin (oral): 400 twice daily | Combination group had a significantly shorter median time from start of study treatment to negative nasopharyngeal swab (7 days [IQR 5-11]) than the LPV/RTV group (12 days [8-15]; hazard ratio 4.37 [95% CI 1.86-10.24], p=0.0010) | AEs included nausea and diarrhoea with no difference between the two groups. One patient in the control group discontinued LPV/RTV because of biochemical hepatitis | RoB 2, some concerns risk of bias | No patients died during the study |
|  |  |  |  | each group were similar |  | PLUS<br>IFN-beta-1b (SCI): three doses of 8 million IUs of interferon beta-1b on alternate days (combination group); for 14 days |  |  |  |  |
| Li et al. 2020 []; China | Randomized blinded trial; single center | Mean (SD), 49.4 (14.7) | 40 (46.5) | Mild/moderate confirmed COVID-19 cases aged 18–80 y<br>Baseline characteristics of the three groups were comparable | 34 patients received:<br>LPV/RTV (oral): 200/50 mg twice daily for 7-14 days | 35 patients received:<br>Umifenovir (oral): 200mg three times daily for 7-14 days<br>OR<br>17 patients received no antiviral therapy | Rate of positive-to-negative conversion of SARS-CoV-2 nucleic acid was similar between groups (all p>0.05)<br>There were no differences between groups in the rates of antipyresis, cough alleviation, or improvement of chest CT at days 7 or 14 (all p>0.05)<br>At day 7, 23.5% patients in the LPV/RTV group, 8.6% in the umifenovir group and 11.8% in the control group showed a deterioration in clinical status from moderate to severe/critical (p=0.206) | Overall, 35.3% patients in the LPV/RTV group and 14.3% in the umifenovir group experienced AEs<br>No apparent AEs occurred in the control group | RoB 2, high risk of bias | Study was blinded to participants, physicians and radiologists who reviewed data but open label to clinicians who recruited patients and research staff<br>All three groups were treated with |

|  |  |  |  |  |  |  |  |  |  | Standard care* if in need |
| --- | --- | --- | --- | --- | --- | --- | --- | --- | --- | --- |
| Lan et al. 2020 []; China | Retrospective; cohort; multicenter | Mean (SD), 55.8 (15.2) | 37 (50.7) | Confirmed COVID-19 cases treated with LPV/RTV alone or combined with umifenovir<br>Different age, sex, and baseline demographics in each group | 34 patients received:<br>LPV/RTV (oral): 400/100 mg twice daily for 14 days | 39 patients received:<br>LPV/RTV (oral): 400/100 mg twice daily<br>PLUS<br>Umifenovir (oral): 200mg three times daily; at least for 3 days | Treatment with LPV/RTV alone was not difference from LPV/RTV combined with umifenovir in overall cure rate (92.3% and 97.1%, respectively)<br>In a modified ITT analysis, LPV/RTV combined with umifenovir led to a median time of hospital stay that was shorter by 1.5 days (12.5 days vs. 14 days)<br>COVID-19 RNA clearance was 92.3% in LPV/RTV and 97.1% in combination therapies group<br>Mean time of virus turning negative was 11.5±9.0 days in combination group compared to 9.9±7.5 in single therapy group | Not reported | NOS, 5 | All eligible patients received standard care* if necessary |
| Wen et al. 2020 []; China | Retrospective; cohort; single center | Mean (SD), 49.9 (16.1) | 81 (45.5) | Confirmed COVID-19 cases aged ≥18 years with hospital stay longer than 14 days<br>No statistically significant difference in baseline characteristics before treatment | 59 patients received:<br>LPV/RTV (oral): 200/50 mg twice daily for 7 days | 36 patients received:<br>Umifenovir (oral): 200mg three times daily for 7 days<br>OR | Time for the new coronavirus nucleic acid in the pharyngeal swab to turn negative was (10.20 ± 3.49 d) in LPV/RTV group, (10.11 ± 4.68 d) in umifenovir group, (10.86 ± 4.74 d) in LPV/RTV plus umifenovir group, and (8.44 ± 3.51 d) in conventional group<br>No significant difference in the rate of nasopharyngeal swab new coronavirus nucleic acid | AEs in the three groups of patients using antiviral drugs was significantly higher than that in the conventional treatment group ($\chi^2=14.875$, p=0.002) | NOS, 5 | All three groups were treated with Standard care* if in need |
| | | | | between patients in LPV/RTV group, umifenovir group, combination (LPV/RTV and umifenovir) group and conventional treatment (no antiviral therapy) group | | 25 patients received:<br>Combined antiviral therapies (LPV/RTV AND umifenovir ; same dosages for 7 days)<br>OR<br>58 patients received no antiviral therapy (conventional group) | conversion, clinical symptom improvement rate, and lung infection imaging improvement rate (p>0.05)<br>There was a statistically significant difference in the ratio of normal/mild to severe/critically severe on the 7th day in the four groups ($\chi^2=9.311$, P=0.017): the combined group (24.0%), umifenovir group (16.7%), LPV/RTV group (5.4%), conventional treatment group (5.2%) | | | |
| Jun et al. 2020 []; China | Retrospective; cohort; single center | Median (IQR), 48 (35-62) | 69 (51.5) | Confirmed COVID-19 cases<br>No statistically significant differences in the demographic data, clinical manifestations, laboratory examinations, and chest CT examination of patients in the LPV/RTV group, | 52 patients received:<br>LPV/RTV (oral): 200/50 mg twice daily for 5 days | 34 patients received:<br>Umifenovir (oral): 200mg three times daily for 5 days<br>OR<br>48 patients received no antiviral therapy | Median time for the body temperature to return to normal in the umifenovir group and the LPV/RTV group was 6 days, and the conventional group was 4 days ($\chi^2= 2.37$, p=0.31)<br>Median time of viral nucleic acid negative in respiratory tract specimens of the three groups was 7 days after treatment<br>Viral nucleic acid negative in the LPV/RTV group was 71.8% and 82.6% in the umifenovir group, | 17.3% in the LPV/RTV group had AEs, including nausea, diarrhea and other GI symptoms; 8.8% in the umifenovir group had AEs, including diarrhea; 8.3% in the control group had AEs such as anorexia and diarrhea ($\chi^2= 2.33$, p=0.33) | NOS, 5 | All patients received IFN α2b spray therapy and standard care* |
|  |  |  |  | umifenovir group, and control (no antiviral therapy) group (all p>0.05) |  | (conventional group) | the conventional group was 77.1% ($\chi^2=0.46$, p=0.79)<br>42.3% patients in the LPV/RTV group, 35.3% patients in the umifenovir group, and 52.1% patients in the conventional group still had progressive imaging on the 7th day after treatment ($\chi^2=2.38$, p=0.30) |  |  |  |
| Yan et al. 2020 []; China | Retrospective; cohort; single center | Median (IQR), 52 (35-63) | 54 (45) | Confirmed COVID-19 cases and had the available RNA viral data to estimate the duration of viral shedding | 78 patients received: LPV/RTV (oral): 200/50 mg twice daily for 10 days or more | 42 patients received no antiviral therapy (conventional group) | Median duration of viral shedding was shorter in the LPV/RTV treatment group than that in no LPV/RTV treatment group (median, 22 days vs. 28.5 days, p=0.02)<br>Patients who started LPV/RTV treatment within 10 days from symptom onset had a shorter duration of SARS-CoV-2 RNA shedding than other patients who began after 10 days (median 19 days vs. 27.5 days, p<0.001) | Not reported | NOS, 5 | Many patients received and standard care* if in need |
| Yuan et al. 2020 []; China | Retrospective; cohort; single center | Median (range), 40 (1-78) | 42 (45) | Confirmed COVID-19 cases of mild and/or moderate symptoms and critical conditions<br>Significant different illness onset on the most common symptoms (fever, fatigue and diarrhoea) | 46 patients received: IFN-α + LPV/RTV (dosages, durations were not reported) | 41 patients received: IFN-α + LPV/RTV PLUS<br>Ribavirin; (dosages, durations were not reported) | No significant difference on average LOS or PCR negative conversion times among different antivirus treatment groups. Correlation analysis indicated that the duration of hospital stay was significantly correlated with PCR negative conversion times in IFN-α + lopinavir/ritonavir + ribavirin group (p=0.0215), as well as IFN-α + lopinavir/ritonavir group (p=0.012) | Not reported | NOS, 6 | Approximately 51% were aged $\leq 40$ y, including 2 children under 3 y |
|  |  |  |  |  |  |  | Average LOS and IFN treatment duration of moderate group were 14.12 (13.34–14.90) days and 14.24 (13.45–15.03) days, respectively, while those of the severe group took average 2.08 days and 1.44 days longer |  |  |  |
| Zhu et al. 2020 []; China | Retrospective; cohort; multicenter | Mean (SD), 39.8 (17.6) | 26 (52) | Confirmed COVID-19 cases<br>No significant difference in age and sex between the two groups | 34 patients received:<br>LPV/RTV (oral): 200/50 mg twice daily for 7 days | 16 patients received:<br>Umifenovir (oral): 200mg three times daily (duration was not reported) | No difference in fever duration between the two groups (p=0.61)<br>On day 14 after the admission, no viral load was detected in umifenovir group, but the viral load was found in 44.1% patients treated with LPV/RTV<br>Patients in the umifenovir group had a shorter duration of positive RNA test compared to those in the LPV/RTV group (p<0.01) | No apparent SEs were found in both groups | NOS, 6 | All patients received and standard care* if in need |
| Ye et al. 2020 []; China | Retrospective; cohort; single center | Range (5-68), of which 9 were <30 and 38 were >30 | 22 (46.8) | Confirmed COVID-19 cases treated with LPV/RTV or not during hospitalization<br>Different age, sex, and baseline demographics in each group | 42 patients received:<br>LPV/RTV (oral): 400/100 mg twice daily or 800/200 mg once daily<br>PLUS<br>Umifenovir (oral): | 5 patients received:<br>Umifenovir (oral): 200mg three times daily<br>PLUS<br>IFN-α1b (aerosol inhalation): | Patients in the LPV/RTV group returned to normal body temperature in a shorter time (test group: 4.8±1.94 days vs. control group: 7.3±1.53 days, p=0.0364)<br>Patients in the LPV/RTV group were able to turn negative in a shorter period of time (LPV/RTV group: 7.8±3.09 days vs. control group: 12.0±0.82 days, p=0.0219) | Increased level of ALT enzyme in the LPV/RTV group | NOS, 5 | All patients received and standard care* if in need |
| | | | | | 200mg three times daily<br>PLUS<br>IFN-α1b (aerosol inhalation):<br>5 million IUs twice daily;<br>(durations of use were not reported) | 5 million IUs twice daily;<br>(durations of use were not reported) | | | | |

Abbreviations: AEs, adverse events; ALT, alanine aminotransferase; COVID-19, coronavirus disease 2019; CT, computed tomography; FiO<sub>2</sub>, fraction of inspired oxygen; FPV, favipiravir; GI, gastrointestinal; IFN, interferon; IQR, interquartile range; ITT, intention-to-treat; IUs, international units; LOS, length of hospital stay; LPV/RTV, lopinavir/ritonavir; NA, not applicable; NEWS2, National Early Warning Score 2; NHC, National Health Commission of China; NOS, Newcastle Ottawa Scale; PaO<sub>2</sub>, partial pressure of oxygen; RoB 2, Version 2 of the Cochrane risk-of-bias tool for randomized trials; ROBINS-I, Risk of bias in non-randomized studies - of interventions; RT-PCR, real-time reverse transcription-polymerase chain reaction; SaO<sub>2</sub>, oxygen saturation; SARS-CoV-2, severe acute respiratory syndrome coronavirus 2; SCI, subcutaneous injection; SEs, side effects
\*Standard care comprised, as necessary, supplemental oxygen, noninvasive and invasive ventilation, antibiotic agents, vasopressor support, renal-replacement therapy, and extracorporeal membrane oxygenation (ECMO).

**Figure 1:**
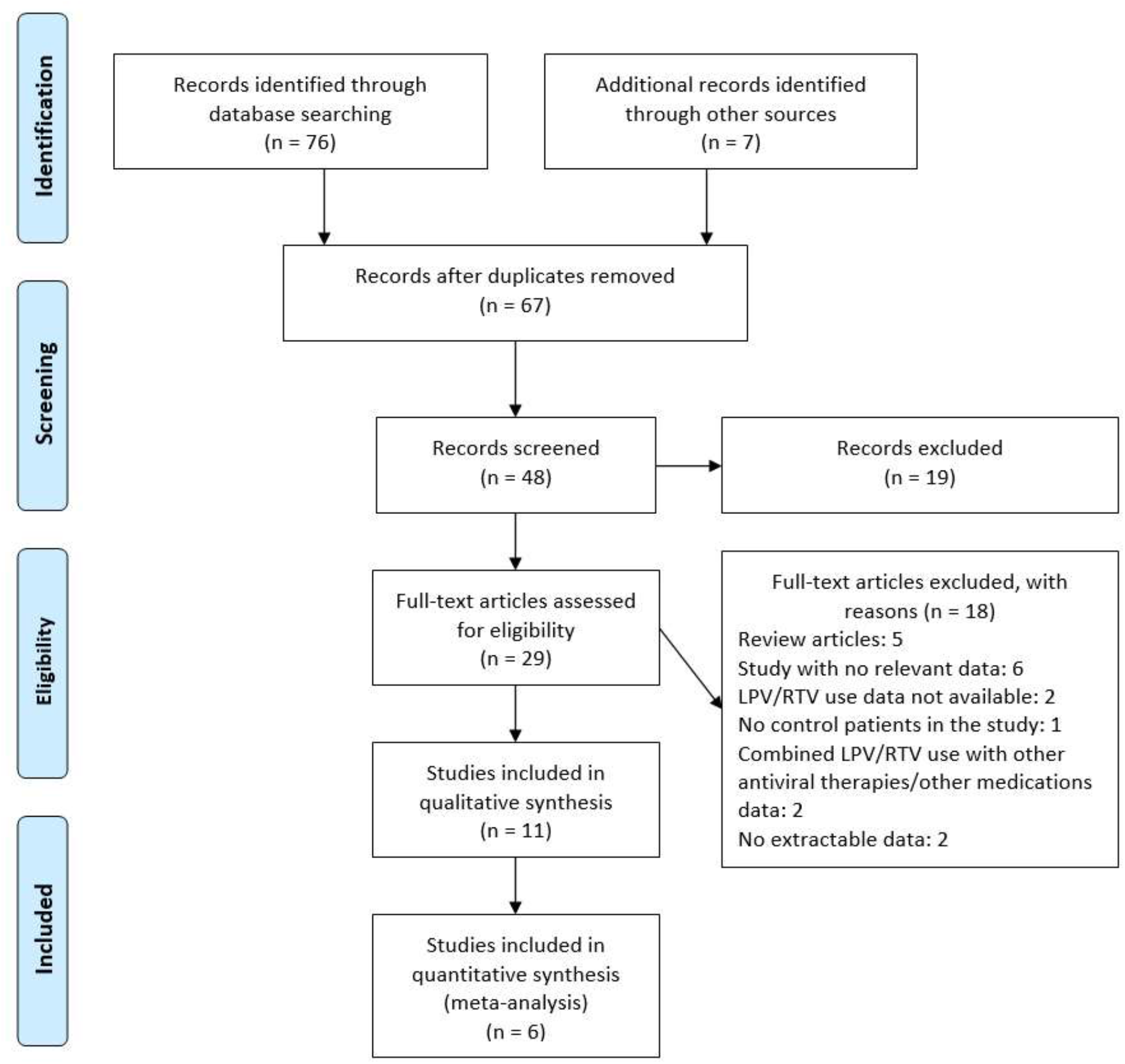
PRISMA flow chart of the included studies. LPV/RTV, lopinavir/ritonavir; PRISMA, Preferred Reporting Items for Systematic reviews and Meta-analysis.

## Comparison 1: efficacy and safety of lopinavir/ritonavir versus no antiviral therapy (conventional therapy) or control

A total of six studies (23-28) reported on lopinavir/ritonavir versus no antiviral therapy (conventional therapy) or control (n=594) in terms of efficacy and safety.

### A. Virological cure on day 7 post initiation of therapy (+ve to −ve PCR: nondetection of SARS-CoV-2 in nasopharyngeal swab)

1. Lopinavir/ritonavir versus no antiviral therapy (conventional cure): virologic cure at day 7 post initiation of therapy: Three studies reported a virologic cure (n = 171 in lopinavir/ritonavir alone arm vs n = 117 in conventional arm) on day 7 (23, 25, 28). Significant mean difference was observed between the two arms in terms of virological cure (mean difference = −0.81 day; 95% CI, −4.44 to 2.81; *P* = 0.007, *I*^*2*^ = 80%) (Figure 1.1).

**Figure 1.1:**
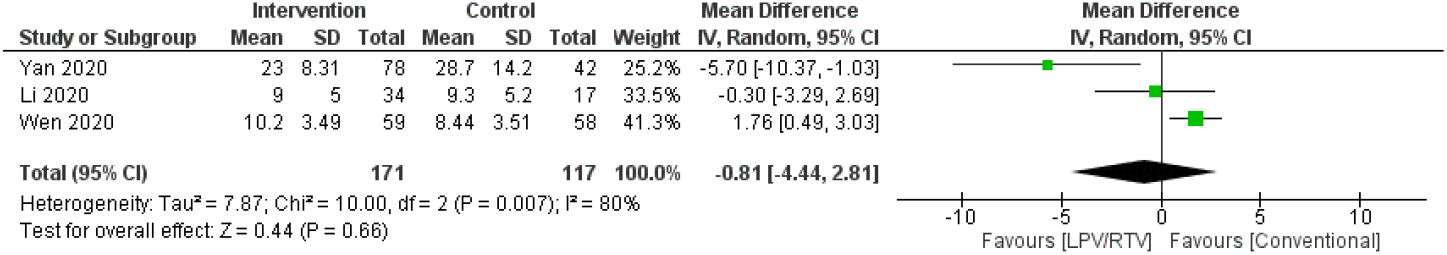
Time from +ve to −ve PCR (days) (LPV/RTV vs no antiviral treatment or conventional). CI, confidence interval; df, degrees of freedom; lopinavir/ritonavir (LPV/RTV)
2. Lopinavir/ritonavir vs umifenovir: Virologic cure at day 7 post initiation of therapy: Three studies reported on virological cure (n = 127 in lopinavir/ritonavir alone arm vs n = 87 in umifenovir arm) on day 7 (23, 26, 28). No significant mean difference was observed between the two arms in terms of virological cure (mean difference = 0.95 day; 95% CI, −1.11 to 3.01; *P* = 0.09, *I*^*2*^ = 58%) (Figure 1.2).

**Figure 1.2:**
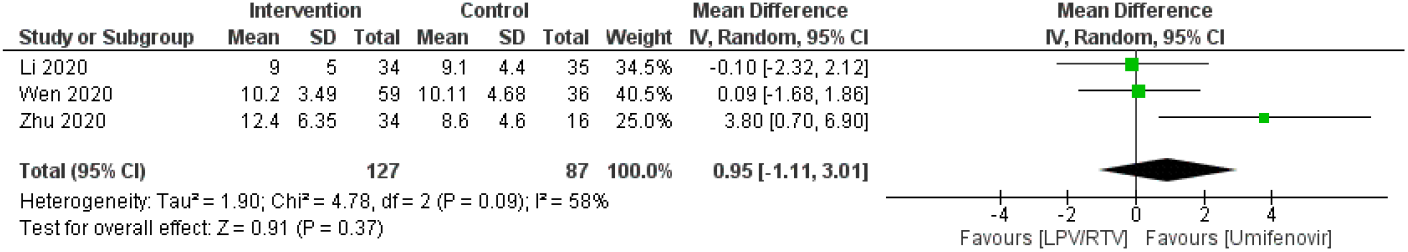
Time from +ve to −ve PCR (days) (LPV/RTV vs umifenovir). CI, confidence interval; df, degrees of freedom; LPV/RTV, lopinavir/ritonavir
3. Lopinavir/ritonavir vs umifenovir plus lopinavir/ritonavir: Virologic cure at day 7 post initiation of therapy: Two studies reported on virological cure (n = 93 in lopinavir/ritonavir alone arm vs n = 75 in umifenovir plus lopinavir/ritonavir arm) on day 7 (24,28). No significant mean difference was observed between the two arms in terms of virological cure (mean difference = −0.83 day; 95% CI, −2.45 to 0.78; *P* = 0.66, *I*^*2*^ = 0%) (Figure 1.3).

**Figure 1.3:**
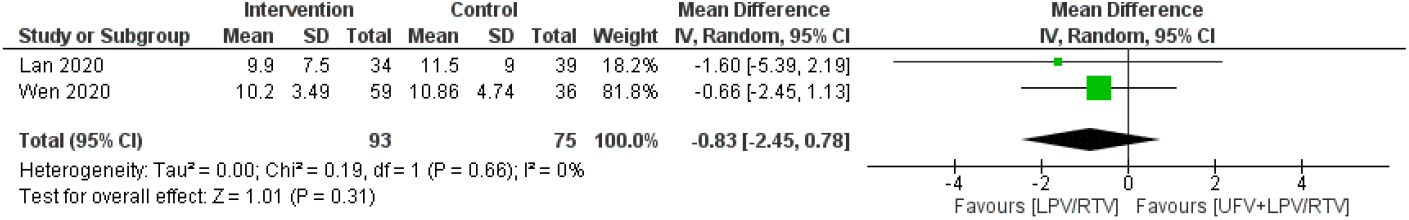
Time from +ve to −ve PCR (days) (LPV/RTV vs LPV/RTV plus umifenovir combination). CI, confidence interval; df, degrees of freedom; LPV/RTV, lopinavir/ritonavir; UFV, umifenovir

### B. Clinical cure (time to body temperature normalization and time to cough relief)

#### 1. Time to body temperature normalization

##### 1.1 Lopinavir/ritonavir vs umifenovir

Two studies reported on time to temperature normalization (n = 93 in lopinavir/ritonavir alone arm vs n = 71 in umifenovir arm) [23, 28]. No significant association was observed between the two arms in terms of temperature normalization (OR = 0.87 day; 95% CI, 0.42 to 1.78; *P* = 0.61, *I*^*2*^ = 0%) (Figure 1.4).

**Figure 1.4:**
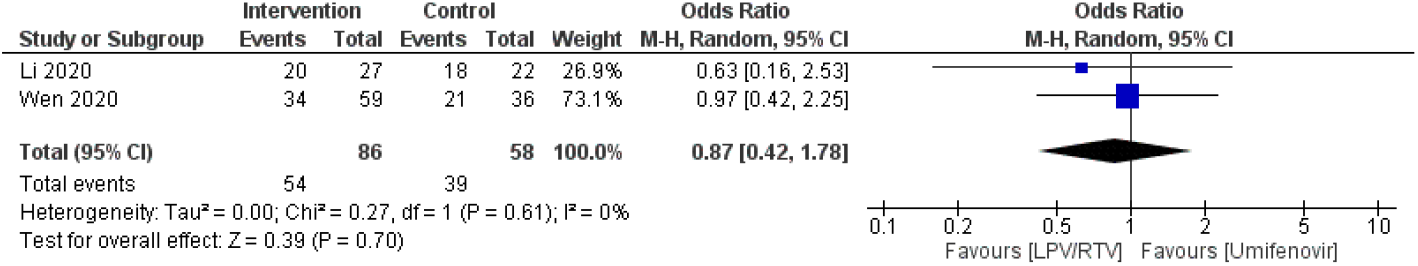
Time to body temperature normalization (days) (LPV/RTV vs umifenovir). CI, confidence interval; df, degrees of freedom; LPV/RTV, lopinavir/ritonavir

##### 1.2 Lopinavir/ritonavir versus no antiviral therapy (conventional)

Two studies reported on time to temperature normalization (n = 93 in lopinavir/ritonavir alone arm vs n = 75 in conventional arm) [23, 28]. No significant association was observed between the two arms in terms of temperature normalization (OR = 0.99 day; 95% CI, 0.49 to 1.99, *P =* 0.35, *I*^*2*^ = 0%) (Figure 1.5).

**Figure 1.5:**
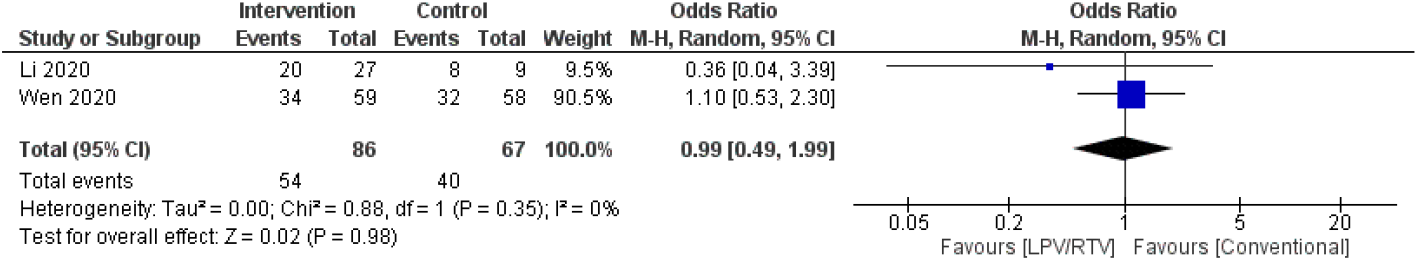
Time to body temperature normalization (days) (LPV/RTV vs no antiviral treatment or conventional). CI, confidence interval; df, degrees of freedom; LPV/RTV, lopinavir/ritonavir

### 2. Duration of cough

#### 2.1 Lopinavir/ritonavir versus umifenovir: Rate of cough alleviation after 7 days of therapy

Two studies reported on cough alleviation (n = 93 in lopinavir/ritonavir alone arm vs n = 71 in umifenovir arm) (23, 28). Lopinavir/ritonavir alone arm had a significant lower number of cough days by 0.62 (95% CI 0.06 to 6.53, *P* = 0.02; *I*^*2*^ = 81%) (Figure 1.6).

**Figure 1.6:**
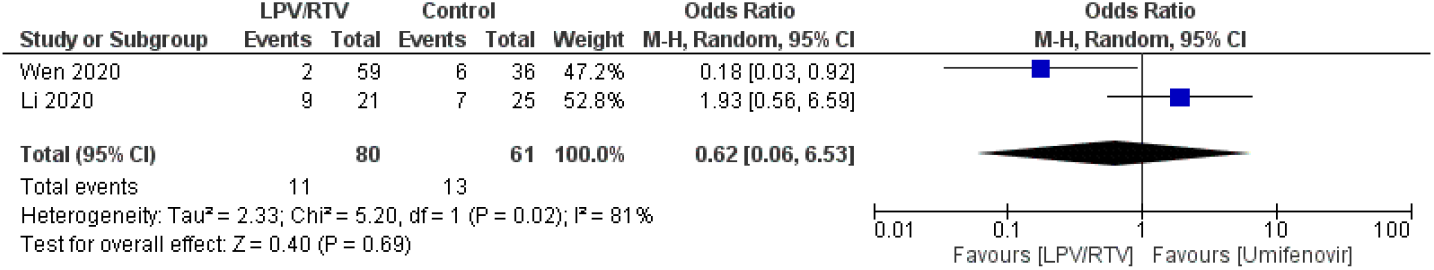
Rate of cough alleviation after 7 days of treatment (LPV/RTV vs umifenovir). CI, confidence interval; df, degrees of freedom; LPV/RTV, lopinavir/ritonavir

#### 2.2 Lopinavir/ritonavir vs no antiviral therapy (conventional): Rate of cough alleviation after 7 days of therapy

Two studies reported on cough alleviation (n = 93 in lopinavir/ritonavir alone arm vs n = 75 in conventional arm) (23, 28). No significant association was observed between the two arms in terms of cough alleviation (OR = 0.87 day; 95% CI, 0.10 to 7.16; *P* = 0.08, *I*^*2*^= 67%) (Figure 1.7).

**Figure 1.7:**
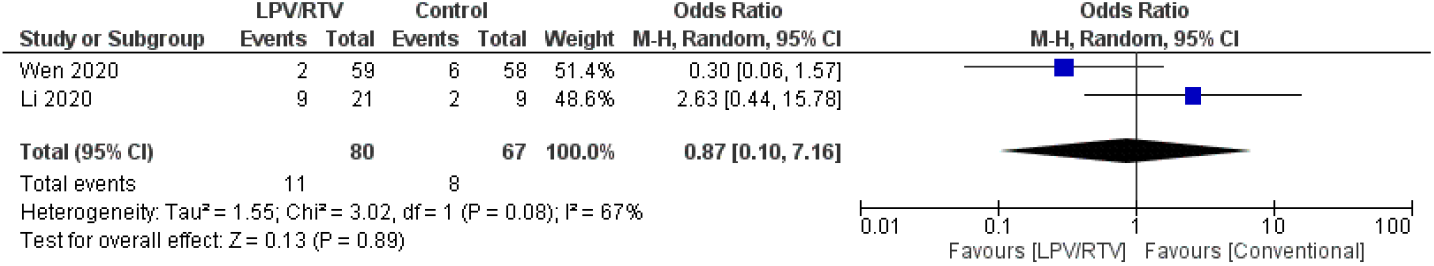
Rate of cough alleviation after 7 days of treatment (LPV/RTV vs no antiviral treatment or conventional). CI, confidence interval; df, degrees of freedom; LPV/RTV, lopinavir/ritonavir

### C. Radiological progression during drug treatment

#### 1 Rate of improvement on chest CT after 7 days of treatment

##### 1.1 Lopinavir/ritonavir vs umifenovir

In terms of CT evidence of radiological progression of pneumonia/lung damage (n = 59 in the lopinavir/ritonavir arm vs n = 71 in umifenovir arm), treatment with lopinavir/ritonavir resulted in no significant decrease in the radiological progression (OR = 0.80; 95% CI, 0.42 to 1.54; *P* = 0.59, *I*^*2*^ = 81%) (Figure 1.8).

**Figure 1.8:**
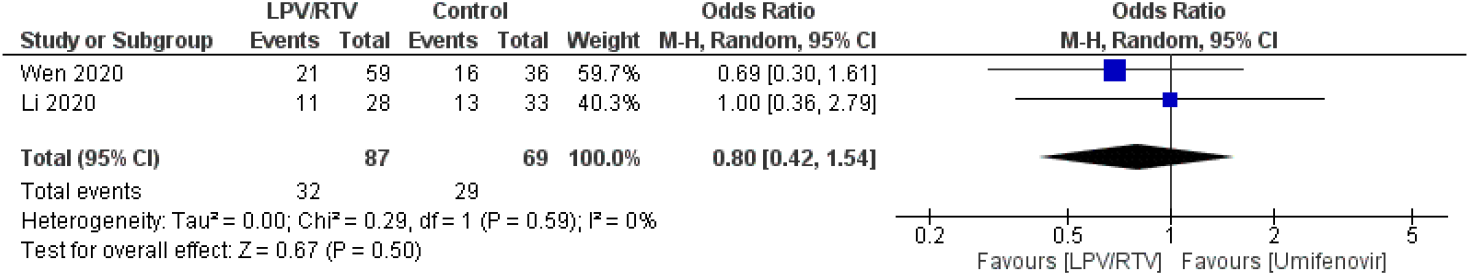
Rate of improvement on chest CT after 7 days of treatment (LPV/RTV vs umifenovir). CI, confidence interval; df, degrees of freedom; LPV/RTV, lopinavir/ritonavir

##### 1.2 Lopinavir/ritonavir vs no antiviral therapy (conventional)

In terms of CT evidence of radiological progression of pneumonia/lung damage (n = 71 in the lopinavir/ritonavir arm vs n = 75 in conventional arm), treatment with lopinavir/ritonavir resulted in no significant decrease in the radiological progression (OR = 0.69; 95% CI, 0.36 to 1.31; *P* = 0.42, *I*^*2*^ = 0%) (Figure 1.9).

**Figure 1.9:**
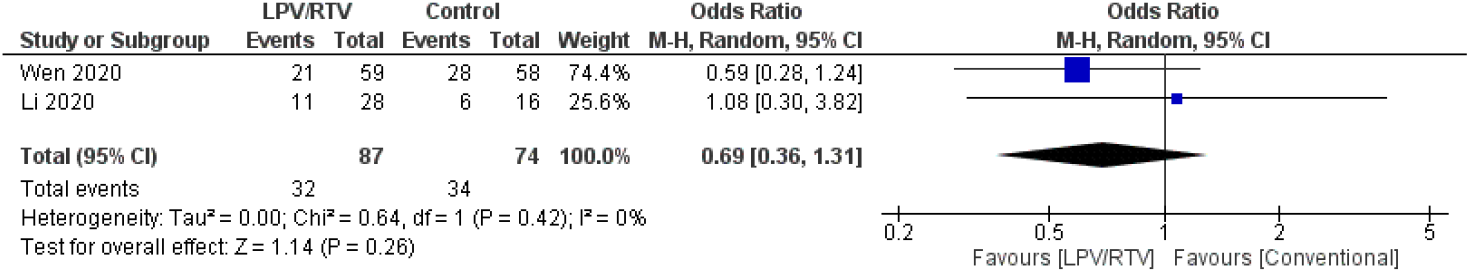
Rate of improvement on chest CT after 7 days of treatment (LPV/RTV vs no antiviral treatment or conventional). CI, confidence interval; df, degrees of freedom; LPV/RTV, lopinavir/ritonavir

### D. Safety and tolerability

#### 1.1 Rate of adverse events of treatment: Lopinavir/ritonavir vs umifenovir

A greater number of adverse events were reported for lopinavir/ritonavir (n=45) relative to the umifenovir arm (n=14) (OR = 2.66; 95% CI, 1.36 to 5.19; *P* = 0.004, *I2* = 0%; Figure 1.10).

**Figure 1.10:**
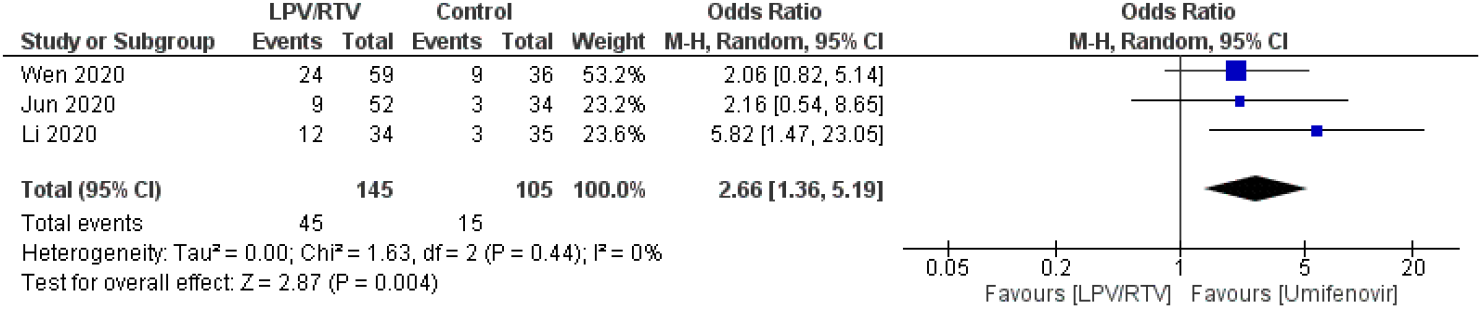
Rate of adverse events of treatment (LPV/RTV vs umifenovir). CI, confidence interval; df, degrees of freedom; LPV/RTV, lopinavir/ritonavir

#### 1.2 Rate of adverse events of treatment: Lopinavir/ritonavir versus no antiviral treatment (conventional)

A greater number of adverse events were reported lopinavir/ritonavir (n=45) than antiviral treatment or conventional (n=10) (*P* = 0.0007) (Figure 1.11).

**Figure 1.11:**
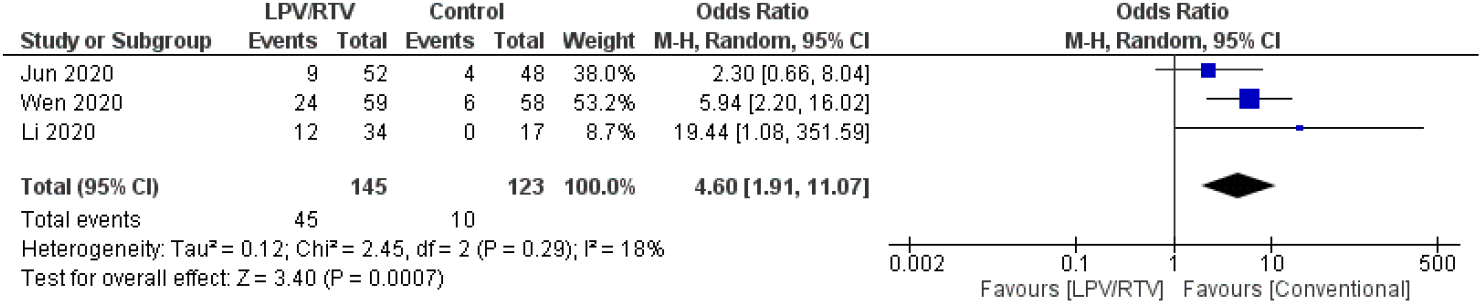
Rate of adverse events of treatment (LPV/RTV vs no antiviral treatment or conventional). CI, confidence interval; df, degrees of freedom; LPV/RTV, lopinavir/ritonavir

## Comparison 2: Efficacy and safety of lopinavir/ritonavir along in combination with other agents versus no antiviral therapy (conventional therapy) or control

A total of four studies evaluated efficacy of LPV/RTV plus interferon (IFN) [29-32] and three studies [29-31] evaluated the safety of the combination. Other studies evaluated the efficacy of LPV/RTV plus standard care [12], ribavirin [30], or umifenovir [31], and evaluated safety of these combinations.

In terms of the efficacy of the combination in patients with COVID-19, LPV/RTV plus IFN combination in addition to ribavirin was safe and superior to LPV/RTV alone by shortening median time from start of study treatment to negative nasopharyngeal swab (7 days [IQR 5–11]) compared to the LPV/RTV arm (12 days [8–15]; hazard ratio 4·37 [95% CI 1·86–10·24], p=0·001) (30). Additionally, combination treatment with LPV/RTV plus IFN and umifenovir had a more evident therapeutic effect in a shorter time by normalizing body temperature (4.8±1.94 days vs. 7.3±1.53 days, p=0.03) and turning PCRs to negative (7.8±3.09 days vs. 12.0±0.82 days, p=0.02) compared to the umifenovir plus IFN arm with no evident toxic and side effects [31]. However, the use of LPV/RTV plus IFN combination resulted in less therapeutic responses on COVID-19 in terms of viral clearance [median (interquartile range, IQR), 4 (2.5–9) d versus 11 (8–13) d, p<0.001) and chest CT changes (91.43% vs 62.22%), p=0.004] compared to the favipiravir plus IFN combination. Favipiravir arm patients had less adverse events (AEs) compared to the LPV/RTV arm (11.43% vs 55.56%) (p<0.001) (29). Additionally, no significant difference on average PCR negative conversion times among IFN plus LPV/RTV or IFN plus LPV/RTV plus ribavirin treatment arms [32].

The combination of LPV/RTV, in addition to standard care, or standard care alone exhibited no difference in the time to clinical improvement (hazard ratio for clinical improvement, 1.31; 95% CI, 0.95 to 1.80) with a similar 28-day mortality (19.2% vs. %; difference, −5.8 percentage points; 95% CI, −17.3 to 5.7).

## Discussion

This systematic review included 11 articles relating to the efficacy and safety of lopinavir/ritonavir in COVID-19 patients, with a total of 1,192 patients included, and only six articles that comprised 594 patients had findings on the efficacy and safety of lopinavir/ritonavir in treatment of COVID-19 verses control/conventional therapy were deemed legible for quantitative synthesis (meta-analysis) [23-28].

In terms of virological cure, three studies reported less time in days for LPV/RTV arm (n = 171) compared with no antiviral therapy (conventional) (n= 117); however, the overall effect was not significant (mean difference = −0.81 day; 95% CI, −4.44 to 2.81; *P* = 0.66), similarly the virological cure for lopinavir/ritonavir alone (n = 127) versus the umifenovir arm (n = 87) (*P* = 0.37), or lopinavir/ritonavir versus umifenovir plus lopinavir/ritonavir (*P* = 0.31) [23-28].

Two studies reported on time to temperature normalization with no significant effect of LPV/RTV (n = 93) versus umifenovir (n = 71) arm), (OR = 0.87 day; 95% CI, 0.42 to 1.78; (*P* = 0.70), I2 = 0%), or alleviation of cough duration (p = 0.69) [23, 28]. The total number of cough days was found to be lower in the LPV/RTV arm compared with the umifenovir arm or no antiviral therapy (conventional) arm after 7 days of treatment; however, the overall effect was found to be not significant [23,28].

Interestingly in this study, treatment with lopinavir/ritonavir (n = 93) versus umifenovir plus lopinavir/ritonavir (n = 75) arm did not reveal any significant mean difference between the two arms in terms of virological cure at day seven. In contrast, a favorable therapeutic effect for umifenovir was observed in a small cohort study when the drug was combined with lopinavir/ritonavir treatment in sixteen COVID-19 patients rather than lopinavir/ritonavir alone (n=17) [33].

In another study that involved 81 COVID-19 patients, the umifenovir treatment group had a longer hospital stay than patients in the control group (13 days (IQR 9–17) vs 11 days (IQR 9–14), p 0.04) [34].

Of note, umifenovir, which is branded as Arbidol®, has a wide antiviral activity against RNA and DNA viruses, and is licensed in Russia and China for treatment and prophylaxis of influenza, is recommended for treatment of MERS-CoV, was investigated in SARS-CoV, and is currently being trialed in COVID-19 patients [35].

In terms of CT evidence of radiological progression of pneumonia/lung damage of lopinavir/ritonavir arm versus umifenovir, although a fewer number of patients exhibited radiological progression in the LPV/RTV arm compared with the umifenovir arm or no antiviral therapy (conventional) arm after 7 days of treatment, this effect was not significant (*P* = 0.59). Similarly, with lopinavir/ritonavir (n=71) versus no antiviral therapy [23, 28].

In terms of safety, this study found greater adverse events reported in lopinavir/ritonavir arm versus no antiviral treatment (conventional) or umifenovir respectively.

Adverse events associated with lopinavir–ritonavir alone or in combination with other medicines were reported in COVID-19 patients, and were typically gastrointestinal (GIT) in nature, including nausea, vomiting, and diarrhea [28]; however, serious GIT ADRs such as acute gastritis and GIT bleeding and acute kidney injury (n=3) were also reported (28). It was reported that most ADRs associated with lopinavir–ritonavir in combined groups of medicines are resolved within three days of drug initiation [29].

To address the efficacy and safety of LPV/RTV combined with other drugs in patients with COVID-9, LPV/RTV plus IFN combination in addition to ribavirin was found superior and more safe than LPV/RTV alone by shortening time to negative nasopharyngeal swab compared to the LPV/RTV arm alone [30]. Additionally, a combined treatment regimen of LPV/RTV plus IFN and umifenovir resulted in a shorter time by normalizing body temperature and turning PCRs to negative compared to the umifenovir plus IFN arm with reasonable safety profile [31]. However, the use of LPV/RTV plus IFN combination resulted in less therapeutic responses on COVID-19 in terms of viral clearance and chest CT changes compared to the favipiravir plus IFN combination. Favipiravir arm patients had less AEs compared to the LPV/RTV arm [29]. Additionally, there was no significant difference in average PCR negative conversion times among IFN plus LPV/RTV or IFN plus LPV/RTV plus ribavirin treatment arms [32]. The combination of LPV/RTV, in addition to standard care, or standard care alone revealed no difference in the time to clinical improvement with a similar 28-day mortality. Gastrointestinal AEs were more common in the LPV/RTV arm, but serious AEs were more common in the standard care arm and treatment was stopped early in 13.8% of patients because of AEs [12].

In a recent systematic review (preprint version) that included 69 studies which included therapeutics for COVID-19, lopinavir/ritonavir was found to be the third therapeutic associated with positive outcomes (54.9%) with less negative outcomes (12.3%) compared to systemic corticosteroids (21.3%), remdesivir (16.9%), moxioxacin (13.4%) and oseltamivir (12.5%) [36]; however, further controlled studies are needed to draw a valid conclusion.

The key limitations of this study were the limited number of clinical studies investigating the efficacy and safety of lopinavir/ritonavir combination with the limited number of participants. Another limitation is inability to perform any type of meta-analysis specifically for the results of efficacy and safety of using lopinavir/ritonavir in combination with other agents versus no antiviral therapy (conventional therapy) or control because of the large methodological differences. Despite these limitations, this systematic review provided valuable insight into the efficacy, safety, and clinical outcomes of lopinavir/ritonavir alone or with other antiviral medications.

## Conclusions

The small number of studies included in this systematic review and meta-analysis study did not reveal any statistically significant advantage in efficacy of lopinavir-ritonavir in COVID-19 patients, over conventional or other antiviral treatments. In terms of safety, this study found greater number of adverse events reported in lopinavir/ritonavir arm versus no antiviral treatment (conventional) or umifenovir arms respectively.

There is a general understanding of the need to conduct large randomized clinical trials to determine the efficacy and safety of lopinavir-ritonavir in the treatment of COVID-19. Ideally, these studies should be double-blinded and conducted in a range of settings.

## Data Availability

The datasets used and/or analysed during the current study are available from the corresponding author on reasonable request.

